# Independent and Joint Associations of Sedentary Behavior and Physical Activity with Cardiometabolic Health Markers in the 1970 British Birth Cohort

**DOI:** 10.1101/19009738

**Authors:** Bo-Huei Huang, Mark Hamer, Sebastien Chastin, Annemarie Koster, Natalie Pearson, Emmanuel Stamatakis

## Abstract

**Objective:** To examine the independent and joint associations thigh-worn accelerometry assessed sedentary time and moderate to vigorous physical activity with cardiometabolic health markers.

**Design:** Cross-sectional study embedded in the age-46 wave an established birth cohort, the 1970 British Birth Cohort.

**Setting:** Population-based sample from Great Britain (England, Scotland, and Wales).

**Methods:** Outcome measures included: body mass index, waist-to-hip ratio, blood pressure, glycated hemoglobin, high-density lipoprotein cholesterol, total cholesterol, triglycerides, and c-reactive protein. Sedentary behavior and other physical activity exposures, recorded by a thigh-worn activPAL3 accelerometry, included: daily sedentary time, breaks in sedentary time, daily time spent in moderate-to-vigorous physical activity. Multiple linear regression analyses, multiple logistic regression analyses, and general linear models were conducted as applicable.

**Results:** 4,634 participants were available for the final analysis. After adjusting for potential confounders and moderate-to-vigorous physical activity, daily sedentary time was positively associated with triglycerides (β=0.052 [0.015, 0.089]) and inversely associated with high-density lipoprotein cholesterol (β=-0.015 [-0.022, -0.010]). Daily prolonged sedentary time (≥ 60 minutes) was positively associated with both glycated hemoglobin and log-transformed c-reactive protein (β=0.240 [0.030, 0.440] and 0.026 [0.007, 0.045], respectively) and inversely associated with systolic blood pressure and high-density lipoprotein cholesterol (β=-0.450 [-0.760, -0.150] and -0.013 [-0.022, -0.003], respectively). After adjusting for potential confounders and daily sedentary time, daily breaks in sedentary time were inversely associated with glycated hemoglobin (β=-0.020 [-0.037, -0.003]), and positively associated with both triglycerides and systolic blood pressure (β=0.006 [0.002, 0.010] and 0.030 [0.002, 0.050], respectively). The joint associations of prolonged sedentary time and moderate-to-vigorous physical activity with the prevalence of diabetes were not statistically significant.

**Conclusion:** Prolonged sedentary time (≥ 60 minutes) and daily breaks in sedentary time were deleteriously associated with glycated hemoglobin, although we found no evidence that there were joint moderate-to-vigorous physical activity and sitting associations.

## Introduction

Cardiovascular disease (CVD) is an escalating major global health issue, and physical inactivity is a core etiological factor (1). Sedentary behavior, defined as low energy expenditure (≤1.5 metabolic equivalents) in a sitting or reclining posture during waking times, has been associated with adverse cardiometabolic health and elevated mortality risk (2–4). Current public health guidelines suggest both increasing physical activity levels and decreasing sedentary time to ameliorate the possible CVD-related health burden (1,5). However, the sedentary behavior guidelines still lack consistent, and more robust scientific evidence is needed to support their efficacy.

Results from a recent meta-analysis (2), based on the nine cohort studies measuring sedentary behavior with self-reported questionnaires, found that, independent of physical activity levels, prolonged sedentary time (over 10 hours/day) was associated with CVD-related mortality and incidence rate. On the contrary, another meta-analysis (6), including 13 questionnaire-based cohort studies, suggested that 1 hour of daily physical activity of moderate to vigorous intensity would eliminate the associations between sedentary time and mortality risk. The inconclusive interaction between sedentary time and physical activity for CVD risk might be, in part, due to measurement error (5). Questionnaires are limited in their capacity to accurately characterize physical activity and sedentary profiles as they inquire about selected aspects of movement and have poor validity (7,8). As a result, studies using objective (device-based) measurements are needed to better understand the joint association of the two exposures with cardiometabolic health.

Aside from total sedentary time, other patterns of sedentary behavior, *e*.*g*., sedentary interruptions/breaks and duration of each bout, can be better explored by studies using accelerometry (4). One recent prospective cohort study (9) suggested a direct association between hip-worn accelerometer assessed sedentary time bout duration and all-cause mortality risk. However, wrist/waist-worn accelerometry could not distinguish the components of stationary behavior, *e*.*g*., working at a standing station or sitting in an office. The measured interruption could be either stand/ambulation changes or sit-stand transitions. Thigh-worn accelerometry has better-established validity and reliability for measuring posture, activity intensity, and ambulatory movement in free-living conditions in adults (10–12). To date, the most extensive cross-sectional study using thigh-worn accelerometers (13) suggested that the odds ratios of metabolic syndrome and type 2 diabetes were higher in individuals with higher sedentary time or lower moderate-to-vigorous physical activity. Another relatively small-scale cross-sectional study (14) found that the number of breaks in sedentary time was beneficially associated with cardiometabolic health markers, *e*.*g*., body mass index, waist circumference, high-density lipoprotein cholesterol (HDL-C), and triglycerides (TG). It also suggested that long average sedentary bout duration would exacerbate the cardiometabolic risks, *e*.*g*., lowering HDL-C and elevating TG (14).

However, to date, no large-scale epidemiological study has utilized thigh-worn accelerometry to investigate the cardiometabolic health potential of physical activity and breaks in sedentary time. The aim of this study, therefore, was to examine the independent and joint associations of sedentary time and physical activity with cardiometabolic health markers using thigh-worn accelerometers in a large population-based cohort.

## Methods

### Sample and Design

Data for the present study were drawn from the age-46 wave of the 1970 British Cohort Study (BCS70). The rationale and sampling methods used in the BCS70 are described in detail elsewhere (15). In brief, BCS70 is an observational prospective population-based cohort study, following the lives of 17,287 people born in a single week of 1970 in England, Scotland, and Wales. In 2016, a new wave at age-46 was conducted, which comprised paper-based questionnaires to collect the self-reported information via interviews during the first home visit. Nurses conducted physical examinations and placed the activity monitor on participants during the following visit. The present study includes cross-sectional analyses from the participants who completed the age-46 wave. All participants gave written informed consent, and the age-46 biomedical survey received ethics from NRES Committee South East Coast - Brighton & Sussex (Ref 15/LO/1446).

### Measurements

#### Demographics and contextual variables

Paper-based questionnaires were used to collect information on educational attainment, self-rated general health, disability/limitations, medication history, smoking, and alcohol consumption. The disability/physical limitation was assessed using the European Statistics on Income and Living Conditions (EU-SILC) (16), and the alcohol intake was evaluated using the Alcohol Use Disorders Identification Test – Primary Care Version (AUDIT-PC) (17).

#### Cardiometabolic health markers

During the home visit, a nurse conducted anthropometric tests, blood pressure measurement, blood sampling, and current medication status records for each participant using standard protocols described in detail in the BCS70 age-46 wave user guide (ref). Anthropometry tests included height, weight, body-fat percentage, and waist/hip circumference. Body Mass Index (BMI) was derived as weight (kg) divided by squared height (m^2^), and waist-to-hip ratio (WHR) was computed as the quotient of waist circumference to hip circumference. Both diastolic (DBP) and systolic (SBP) blood pressure were accessed via triplicate measurements. A nurse collected non-fasting blood samples from consenting participants for analyzing glycated hemoglobin (HbA1c), high-density lipoprotein cholesterol (HDL-C), total cholesterol (TC), triglycerides (TG), and c-reactive protein (CRP).

#### Sedentary behavior and other physical activity variables

Sedentary time and physical activity were measured using a thigh-worn activPAL3 monitor (PAL Technologies, Glasgow, UK) according to the protocol for large scale studies defined by Dall et al. to ensure compliance with thigh worn devices (18). The device is a triaxial accelerometry that provides estimated body posture (sitting/reclining/lying, standing) and stepping speed (cadence) based on acceleration information with a sampling frequency of 20 Hz. The wear protocol has been described in detail elsewhere (10). Briefly, at the end of a nurse home visit, a nurse waterproofed and fitted the device on the anterior midline of a participant’s right thigh as recommended by the manufacturer. Participants were asked to wear the device for seven consecutive days without removing it at any time. Participants were asked to return the device using self-addressed envelopes provided by the nurses. After the device was returned, data was downloaded and processed through activPAL3 software and a previously validated open-access program that quantifies non-wear waking times (https://github.com/UOL-COLS/ProcessingPAL/releases/tag/V1.0) (19). The first-day of data were excluded, and subsequent days were defined as the 24 hours between consecutive midnights. Only participants providing at least one valid day, defined as waking wear time more than 10 hours per day, were included for further analysis (13).

### Data Handling

#### Sedentary behavior and physical activity variables

Daily sedentary time (classified as bouts lasting <30 min, 30-60 min, and >60 min), and breaks in sedentary time (number of sit-stand transitions) were calculated using the algorithm program mentioned above (18,19). In the absence of established standards to define prolonged sedentary bouts, both ≥ 60 minutes and ≥ 30 minutes of continuous sedentary time were used as a cutoff (14). Daily time spent in PA of moderate-to-vigorous physical activity (MVPA) was computed using the previously validated program based on cadence (18,20). To compare the demographics and contextual characteristics, participants were divided into three groups based on the tertiles of daily sedentary time. To perform joint association analysis, both daily sedentary time and MVPA time were dichotomized into high and low groups based on median cut points for each behavior. The resulting joint variable comprised four categories of sedentary time and MVPA.

#### Cardiometabolic health markers

CRP was log-transformed as the variable displayed a skewed distribution (21). We calculated the total-to-HDL cholesterol ratio by dividing TC to HDL-C since this ratio has been shown to reflect more informative lipid profile and more predictive of cardiovascular risk (22). A constant was added to the variable for those on medicine (22), *i*.*e*., on lipid-lowering drugs (+25% for TC; −5% for HDL-C; 18% for TG), on BP-lowing drugs (+10 mmHg for DBP and SBP, respectively), and on oral medication for type 2 diabetes (+3.2% for HbA1c).

#### Potential confounders

Sex, education, total waking wear time, self-rated general health, disability/limitations, smoking, and alcohol consumption were used to adjust the statistic models as potential confounders (23). Medication uses were corrected by calculation mentioned above.

### Statistical Analyses

We defined continuous exposures in the present study as sedentary time, breaks in sedentary time, and intensity-specific PA time, with categorical exposures as two sedentary time groups and two MVPA time groups based on each median, while continuous outcomes as BMI (kg/m2), body fat (%), WHR, HbA1c (% of total hemoglobin), TC (mmol/L), HDL-C (mmol/L), TG (mmol/L), CRP (mg/L), SBP (mmHg), total-to-HDL cholesterol ratio, while binary outcomes included hypertension (identified from physician diagnosis and/or SBP ≥ 130 mmHg and/or DBP ≥ 80 mmHg) and diabetes (identified from physician diagnosis and/or HbA1C ≥ 6.5%).

All tests were performed using SAS 9.4 software (SAS Institute, Cary, NC, USA) and were two-sided. Data was shown in mean with either standard deviation or 95% confidence interval as applicable. To examine the differences of demographics and contextual characteristics between tertiles of daily sedentary time, and chi-square tests, ANOVA, and Kruskal-Wallis tests were conducted as applicable. Multiple linear regression analyses were used for continuous exposures and outcomes. Multiple logistic regression analyses were utilized for continuous exposures and binary outcomes. General linear models were performed for independent/joint categorical exposures and continuous outcomes with Bonferroni-corrected pairwise comparisons.

## Results

The age-46 wave recruited 12,368 participants from BCS70 cohort members for either the dress rehearsal or the main stage and collected 8,581 productive cases after interviews (Fig. 1). Among those, 7,888 completed home visits, and 6,060 consented to the enrolment of the accelerometry-based measurement. Eight hundred and sixty-two participants were excluded due to unusable data, and another 560 were excluded because of missing covariates data. A total number of 4,634 participants were available for further analysis prior to biomedical data exclusions. **Table 1** presents the characteristics of the sample by tertiles of daily sedentary time. Male, smoking, degree-educated, poor health self-rated, disability, obesity, type 2 diabetes, and hypertension were characteristics associated with higher daily sedentary time.

**Table 1.** Sample characteristics distribution by daily sedentary time

|  | Tertiles of daily sedentary time (hr/d) |  |  |
| --- | --- | --- | --- |
|  | Low (<8.4) | Medium (8.4-10.1) | High (>10.1) |
| n | 1583 | 1525 | 1526 |
| Age (yrs) | 46.8 (0.7)<sup>a</sup> | 46.8 (0.7) | 46.8 (0.7) |
| Men (%) | 40.6 | 46.4 | 55.3 |
| Smokers (%) | 13.1 | 12.7 | 15.7 |
| Degree educated (%) | 18.7 | 25.7 | 26.7 |
| Poor self-rated health (%) | 1.8 | 3.6 | 6.6 |
| Disability (%) | 2.3 | 5.4 | 7.6 |
| Obese ($\geq 30$ kg/m<sup>2</sup>) | 27.5 | 30.3 | 37.6 |
| Diabetes (%) | 2.7 | 2.4 | 5.7 |
| Hypertension (%) | 6.6 | 6.2 | 8.9 |
| MVPA (hr/d) | 1.4 (0.5) | 1.2 (0.4) | 1.0 (0.4) |
| Device wear days | 6.1 (1.7) | 6.3 (1.4) | 6.1 (1.6) |

<sup>a</sup>Values are in mean (SD) if applicable.

**Figure 1.**
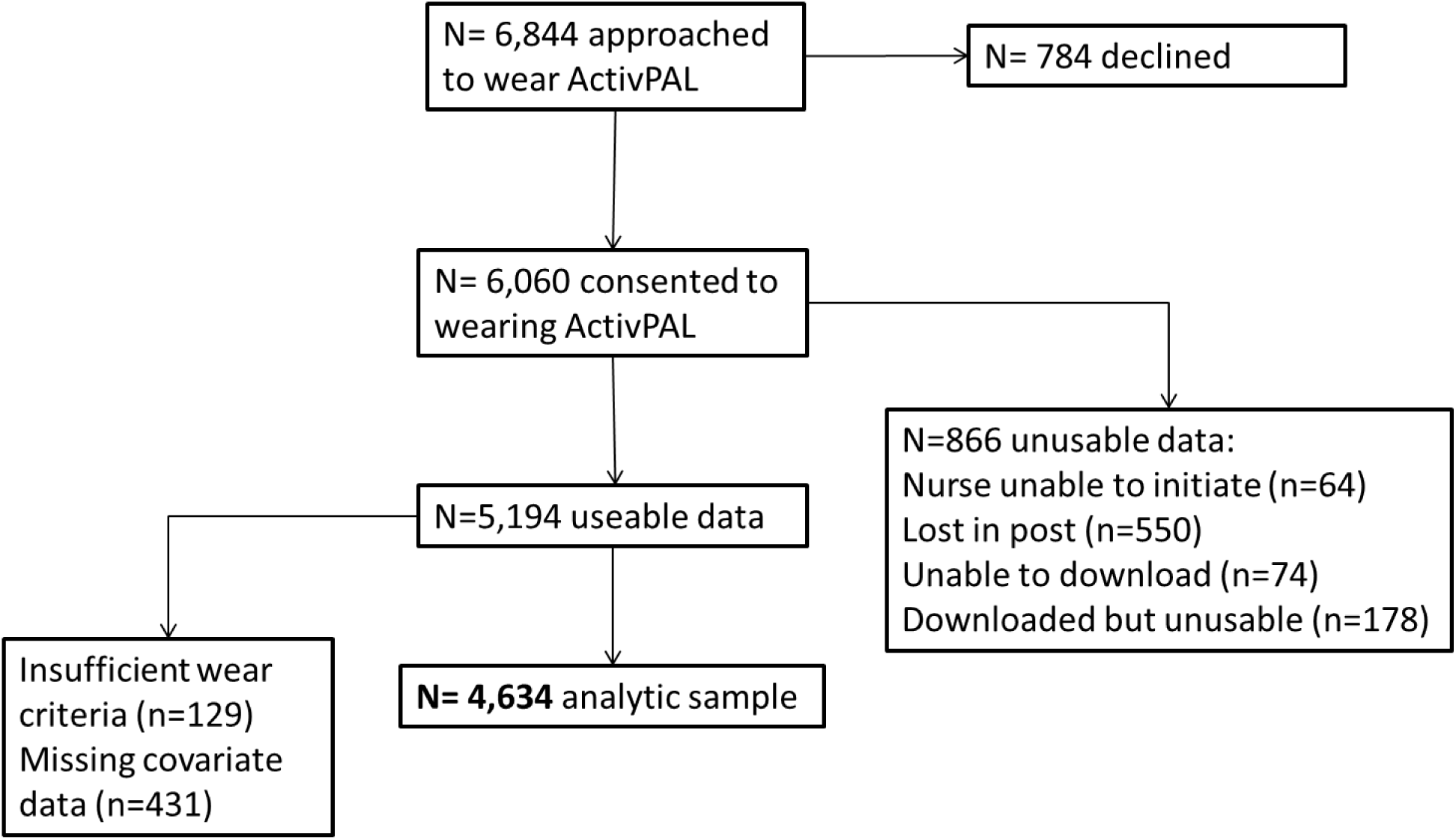
Participants recruitment flowchart

### Association between cardiometabolic health markers and daily sedentary time

As **Table 2** shows, after adjusting for potential confounders and MVPA, daily sedentary time was positively associated with TG (β=0.052 [0.015, 0.089]) but inversely associated with HDL-C (β=-0.015 [-0.022, -0.010]). Daily sedentary time was not significantly associated with HbA1c, log CRP, and SBP. However, when considering daily prolonged sedentary time (≥ 60 minutes) as an independent variable, prolonged sedentary time was positively associated with both HbA1c and log CRP (β=0.240 [0.030, 0.440] and 0.026 [0.007, 0.045], respectively) but inversely associated with SBP and HDL-C (β=-0.450 [-0.760, - 0.150] and -0.013 [-0.022, -0.003], respectively).

**Table 2.** Multivariable association between daily sedentary time (hr/d) and cardiometabolic risk markers

| Risk factor<sup>a</sup> | Model 1<sup>c</sup> | Model 2<sup>d</sup> | Model 3<sup>e</sup> |
| --- | --- | --- | --- |
| **HbA1c (mmol/mol)**<br>(n=3,833) |  |  |  |
| Total sedentary time | 0.31 (0.16, 0.46)<sup>b</sup> | 0.074 (-0.070, 0.22) | 0.074 (-0.091, 0.24) |
| Total prolonged sedentary time | 0.55 (0.36, 0.74) | 0.22 (0.03, 0.41) | 0.24 (0.03, 0.44) |
| **HDL-C (mmol/L)**<br>(n=3,922) |  |  |  |
| Total sedentary time | -0.036 (-0.043, -0.029) | -0.022 (-0.029, -0.016) | -0.015 (-0.022, -0.010) |
| Total prolonged sedentary time | -0.040 (-0.049, -0.031) | -0.021 (-0.030, -0.013) | -0.013 (-0.022, -0.003) |
| **Triglycerides (mmol/L)**<br>(n=2,251) |  |  |  |
| Total sedentary time | 0.055 (0.023, 0.087) | 0.037 (0.004, 0.069) | 0.052 (0.015, 0.089) |
| Total prolonged sedentary time | 0.024 (-0.019, 0.066) | -0.004 (-0.048, 0.039) | -0.002 (-0.048, 0.045) |
| **Log CRP**<br>(n=2,226) |  |  |  |
| Total sedentary time | 0.036 (0.022, 0.050) | 0.013 (0.00, 0.026) | 0.009 (-0.007, 0.024) |
| Total prolonged sedentary time | 0.063 (0.044, 0.081) | 0.029 (0.011, 0.046) | 0.026 (0.007, 0.045) |
| **Systolic BP (mmHg)**<br>(n=4,589) |  |  |  |
| Total sedentary time | 0.25 (0.02, 0.47) | -0.03 (-0.25, 0.19) | -0.10 (-0.35, 0.15) |
| Total prolonged sedentary time | 0.13 (-0.17, 0.42) | -0.36 (-0.64, 0.07) | -0.45 (-0.76, -0.15) |

<sup>a</sup>A constant was added to the variable for those on medicine, *i.e.*, on lipid-lowering drugs (+25% for TC; -5% for HDL-C; 18% for TG), on BP-lowering drugs (+10 mmHg for DBP and SBP, respectively), and on oral medication for type 2 diabetes (+3.2% for HbA1c).
<sup>b</sup>Values are in $\beta$ coefficient and 95% confidence interval.
<sup>c</sup>Adjusted for sex, education, total waking wear time.
<sup>d</sup>Additionally adjusted for smoking, self-rated health, disability, BMI
<sup>e</sup>Additionally adjusted for MVPA.

### Association between daily sit-stand transitions and cardiometabolic health markers

**Table 3** presents the results of the regression models investigating the association between daily sit-stand transitions and cardiometabolic health markers. After adjusting for potential confounders, daily sit-stand transitions were inversely associated with HbA1c (β=-0.020 [-0.037, -0.003]) but positively associated with both TG and SBP (β=0.006 [0.002, 0.010] and 0.030 [0.002, 0.050], respectively). These associations persisted after further adjusting for total daily sitting time. However, there was no association of daily sit-stand transitions with HDL-C and log CRP once the models were adjusted for all confounders.

**Table 3.** Multivariable association between daily sit-stand transitions (times/d) and cardiometabolic risk markers

| Risk factor<sup>a</sup> | Model 1<sup>c</sup> | Model 2<sup>d</sup> | Model 3<sup>e</sup> |
| --- | --- | --- | --- |
| HbA1c (mmol/mol)<br>(n=3,833) | -0.044 (-0.061, -0.027)<sup>b</sup> | -0.020 (-0.037, -0.003) | -0.020 (-0.037, -0.003) |
| HDL-C (mmol/L)<br>(n=3,922) | 0.002 (0.001, 0.003) | 0.000 (-0.001, 0.001) | 0.000 (-0.001, 0.001) |
| Triglycerides (mmol/L)<br>(n=2,251) | 0.003 (-0.001, 0.007) | 0.006 (0.002, 0.010) | 0.006 (0.002, 0.010) |
| Log CRP<br>(n=2,226) | -0.003 (-0.005, -0.002) | -0.001 (-0.002, 0.01) | -0.001 (-0.002, 0.01) |
| Systolic BP (mmHg)<br>(n=4,589) | -0.02 (-0.05, 0.002) | 0.03 (0.002, 0.05) | 0.03 (0.002, 0.05) |

<sup>a</sup>A constant was added to the variable for those on medicine, *i.e.*, on lipid-lowering drugs (+25% for TC; -5% for HDL-C; 18% for TG), on BP-lowering drugs (+10 mmHg for DBP and SBP, respectively), and on oral medication for type 2 diabetes (+3.2% for HbA1c).
<sup>b</sup>Values are in $\beta$ coefficient and 95% confidence interval.
<sup>c</sup>Adjusted for sex, education, total waking wear time.
<sup>d</sup>Additionally adjusted for smoking, self-rated health, disability, BMI
<sup>e</sup>Additionally adjusted for total sitting time.

### Joint association of total daily sedentary time in prolonged bouts and moderate-to-vigorous physical activity with the prevalence of diabetes

**Table 4** shows the odds ratios of the presence of diabetes between different exposure combinations of total daily sedentary time in prolonged bouts and MVPA. After adjustment for potential confounders, there was no significant joint association of prolonged sedentary time and MVPA with the prevalence of diabetes.

**Table 4.** Joint association of total daily sedentary time in prolonged bouts and moderate-to-vigorous physical activity with prevalence of diabetes.

| MVPA<sup>b</sup> | Sedentary time<sup>c</sup><br>in prolonged bout | Cases<sup>d</sup> /N | Model 1<sup>e</sup> | Model 2<sup>f</sup> |
| --- | --- | --- | --- | --- |
| High | Low | 38/1678 | 1.0 (Ref) | 1.0 (Ref) |
| High | High | 36/1075 | 1.43 (0.90, 2.29)<sup>a</sup> | 1.24 (0.76, 2.02) |
| Low | Low | 22/643 | 1.63 (0.95, 2.78) | 1.03 (0.59, 1.81) |
| Low | High | 66/1189 | 2.59 (1.72, 3.89) | 1.24 (0.79, 1.93) |

<sup>a</sup>Values are in odds ratio and 95% confidence interval.
<sup>b</sup>High and low MVPA reflects median cut (~1hr/d).
<sup>c</sup>High and low sitting reflects median cut (~2hr/d).
<sup>d</sup>Diabetes identified from physician diagnosis and/or HbA1C ≥ 48 mmol/mol.
<sup>e</sup>Adjusted for sex, education, total waking wear time.
<sup>f</sup>Additionally adjusted for smoking, self-rated health, disability, BMI

## Discussion

To our knowledge, this is the most extensive and largest cross-sectional study using thigh-worn accelerometry to investigate the joint and independent associations of sedentary time and physical activity with key markers of cardiometabolic health in an established population-based cohort. Using a novel postural allocation technique, we found that prolonged sedentary time was adversely associated with various biomarkers.

### Different sedentary patterns and their associations with cardiometabolic health

Although the adverse health effect of sedentary time has been widely suggested, the roles of different sedentary patterns are still controversial. Our findings suggested that total sedentary time was adversely associated with TG and HDL-C but not associated with other selected cardiometabolic health markers. Inconsistent with this study, systematic reviews of cross-sectional studies using wrist/waist-worn accelerometry have shown discrepant results of associations of total sedentary time with HDL-C and blood glucose despite conclusive adverse associations with insulin and TG (24,25). However, systematic reviews and meta-analyses based on prospective cohort studies showed harmonious results on the detrimental associations of total sedentary time with type 2 diabetes, all-cause mortality, and CVD-related mortality (26–28). These inconclusive results could not only indicate that each measurement method has the different capability of sedentary time estimation, but also imply the importance of understanding different sedentary patterns.

Our results further highlighted the importance of the accumulation of prolonged sedentary time, since only time in prolonged bouts, but not total sedentary time was associated with HbA1c, SBP, and CRP. Further, to our knowledge, the present study is the first observational study, which is in line with the laboratory result, *i*.*e*., breaks in sedentary time were positively associated with glycemic control (29). While previous studies using wrist/waist-worn accelerometry generally suggested that the associations between prolonged sedentary time and cardiometabolic health markers, *e*.*g*., HDL-C, would be independent of total sedentary time (30–34), cross-sectional studies using thigh-worn accelerometry showed discordant results of the prolonged sedentary bouts (14,35). Since wrist/waist-worn measurement could not distinguish different stationary behaviors (5), existing concordant results could arise from pooling both sitting and standing as a single exposure. The definition of a prolonged sedentary bout in the present study was accumulation of a sedentary posture continuously for over 60 minutes. Other studies using less prolonged periods, such as 30 minutes (14), found no associations of prolonged sedentary time with both BP and HbA1c. On the other hand, van der Berg et al. (12) used the 30-min cutoff but defined the number of prolonged boats as the exposure, concluding that the prolonged sedentary bouts were not associated with either the metabolic syndrome or glucose metabolism. These discordant results might indicate that further research is needed to determine a specific cutoff and to investigate the potential dose-response relationship (36).

### Odds for diabetes within daily prolonged sedentary time x weekly MVPA time subgroups

Our study showed no significant joint association of prolonged sedentary time and MVPA with the odds of diabetes. This is in contrast to the only other study investigating a similar hypothesis using thigh-worn accelerometry (13); van der Velde et al. suggested that the risk for type 2 diabetes and the metabolic syndrome was highest in people with low cardiorespiratory fitness combined with either high sedentary time or low MVPA. Similarly, a meta-analysis of questionnaire-based prospective cohort studies suggested that a lower level of PA would exacerbate the disadvantageous associations between sedentary time and hazard ratios of outcomes like all-cause mortality (27). The differences between our study and the two studies mentioned above might come from two different characteristics of participants. Firstly, average daily MVPA time in the present study (∼1.2 hr/d) greatly exceeded the physical activity recommendation (37) and the mean MVPA time of the most active subgroups in both previous studies (13,27). The potential joint association with diabetes risk could be thereby attenuated in the present study. Secondly, the BCS70 age-46 cohort has a low prevalence of diabetes, while the Maastricht study oversampled for participants with type 2 diabetes. The difference between population bases needs to be considered when interpreting the differing results. To conclude, the present study suggested that the odds for diabetes was not different between daily prolonged sedentary time x weekly MVPA time subgroups when participants generally were very physically active.

### Strengths and limitations

A major strength is that our study used thigh-worn accelerometry to continuously record sedentary time and physical activity in free-living conditions, and allows distinction between postures that other devices do not (10–12). Another strength is that our sample was from a representative population-based cohort, increasing the ecological validity of our results. The main limitation of the present study is the cross-sectional study design. However, previous intervention studies have demonstrated consistent results that uninterrupted sedentary time results in adverse changes of cardiometabolic health markers (38), and prospective epidemiological studies have also shown strong evidence for relationships between sedentary time and all-cause/CVD-related mortality (28). The direction could be potentially toward cardiometabolic health outcomes from sedentary exposures. The other potential limitation is residual confounding (13,36), *e*.*g*., CVD history, diet, and light PA, so our results might suffer from potential bias. Finally, all our exposures were compositional data, and our analysis did not account for this (39).

### Conclusion

To the best of our knowledge, this is the largest cross-sectional study using a novel postural allocation technique to objectively measure different patterns of sedentary time and physical activity in free-living settings. Our results suggested that only prolonged sedentary time, but not total sedentary time, was adversely associated with diabetes diagnosis markers, *i*.*e*., HbA1c. However, there were no significant differences in the odds for the presence of diabetes between participants with different prolonged sedentary time and MVPA combinations. Further studies should be conducted to explore the optimized cutoff of a prolonged sedentary bout, and to elucidate potential causality and the dose-response relationship. To sum up, the present study sheds light on the potential relationship between free-living behavior and cardiometabolic health, providing quantitative evidence to both future longitudinal studies and the development of public health guidelines.

## Data Availability

Data for the present study were drawn from the 1970 British Cohort Study and were available on the official website.

https://cls.ucl.ac.uk/cls-studies/1970-british-cohort-study/

